# Patient-specific Quality Assurance Failure Prediction with Deep Tabular Models

**DOI:** 10.1101/2022.10.02.22280624

**Authors:** R. Levin, A. Y. Aravkin, M. Kim

**Affiliations:** University of Washington, Seattle WA

## Abstract

**Background:** Patient-specific quality assurance (PSQA) is part of the standard practice to ensure that a patient receives the dose from intensity-modulated radiotherapy (IMRT) beams as planned in the treatment planning system (TPS). PSQA failures can cause a delay in patient care and increase workload and stress of staff members. A large body of previous work for PSQA failure prediction focuses on non-learned plan complexity measures. Another prominent line of work uses machine learning methods, often in conjunction with feature engineering. Currently, there are no machine learning solutions which work directly with multi-leaf collimator (MLC) leaf positions, providing an opportunity to improve leaf sequencing algorithms using these techniques.

**Purpose:** To improve patient safety and work efficiency, we develop a tabular transformer model based directly on the MLC leaf positions (without any feature engineering) to predict IMRT PSQA failure. This neural model provides an end-to-end differentiable map from MLC leaf positions to the probability of PSQA plan failure, which could be useful for regularizing gradient-based leaf sequencing optimization algorithms and generating a plan that is more likely to pass PSQA.

**Method:** We retrospectively collected DICOM RT PLAN files of 968 patient plans treated with volumetric arc therapy. We construct a beam-level tabular dataset with 1873 beams as samples and MLC leaf positions as features. We train an attention-based neural network FT-Transformer to predict the ArcCheck-based PSQA gamma pass rates. In addition to the regression task, we evaluate the model in the binary classification context predicting the pass or fail of PSQA. The performance was compared to the results of the two leading tree ensemble methods (CatBoost and XGBoost) and a non-learned method based on mean MLC gap.

**Results:** The FT-Transformer model achieves 1.44% Mean Absolute Error (MAE) in the regression task of the gamma pass rate prediction and performs on par with XGBoost (1.53 % MAE) and CatBoost (1.40 % MAE). In the binary classification task of PSQA failure prediction, FT-Transformer achieves 0.85 ROC AUC (with CatBoost and XGBoost achieving 0.87 ROC AUC and the mean-MLC-gap complexity metric achieving 0.72 ROC AUC). Moreover, FT-Transformer, CatBoost, and XGBoost all achieve 80% true positive rate while keeping the false positive rate under 20%.

**Conclusions:** We demonstrate that reliable PSQA failure predictors can be successfully developed based solely on MLC leaf positions. Our FT-Transformer neural network can reduce the need for patient rescheduling due to PSQA failures by 80% while sending only 20% of plans that would not have failed the PSQA for replanning. FT-Transformer achieves comparable performance with the leading tree ensemble methods while having an additional benefit of providing an end-to-end differentiable map from MLC leaf positions to the probability of PSQA failure.

## I. Introduction

Intensity-modulated radiation therapy (IMRT)^1^ achieves a dose distribution that is highly conformal to the target while minimizing the dose to normal tissue by modulating beam intensities within the radiation fields, often termed fluence maps. The beam modulation is performed using multi-leaf collimators (MLC) located within the gantry of a linear accelerator by varying the speed and position of each leaf and gantry angle.

Leaf sequencing algorithms^2,3,4,5,6,7,8^ in the treatment planning system (TPS) optimize the MLC movements to deliver a desirable dose distribution as a treatment planer specifies. Ultimately, final dose distributions to patients are computed using the optimal leaf sequences.

IMRT delivery is a complex, multi-step process with a number of possible sources of noise ranging from computational approximations in the underlying algorithms to physical effects in the linear accelerator components. Therefore, an extensive quality assurance (QA) process is required to prevent any unintended error from reaching the patient and affecting the patient’s clinical outcome. It is current practice in many clinics to perform a patient-specific QA (PSQA) for each patient’s radiation treatment plan^9,10,11^ to ensure that the linear accelerator delivers the correct dose distributions as designed and shown by TPS.

One of the prevalent ways to perform PSQA is using a 3D phantom with an embedded array of detectors to measure the dose delivered using the patient’s treatment beams. Then the computed dose distribution in the TPS is compared with the measured dose distribution, and a gamma analysis is performed to quantify the agreement between the two^12,13^. Sometimes, PSQA fails due to a poor agreement between the computed and measured dose distributions requiring a replanning process and another PSQA, which is often done outside clinic hours. PSQA failure can cause increased workloads and stress for hospital staff members, delay patient treatment, or compromise patient safety if the work has to be rushed to preserve the patient’s original treatment schedule.

To mitigate those issues and improve patient safety, many studies explored PSQA failure prediction. An extensive line of research focused on developing non-learned treatment plan complexity metrics such as modulation complexity score, mean aperture displacement, or small aperture score and investigating their correlation with PSQA failure ^14,15,16,17,18,19,20^. A large number of papers further extended these approaches by developing classical machine learning and deep learning models to predict the PSQA failure based on a vast array of the plan complexity metrics as well as other heuristic features^21,22,23,24,25,26,27,28^. Thongsawad et al. used MLC texture analysis and boosting algorithms for predicting gamma evaluation results^29^. Kimura *et al*. and Huang *et al*. used target metrics alternative to gamma pass rates, such as dose difference^30,31^. Other works leveraged convolutional neural networks to predict the PSQA failure directly from fluence maps^32^ or dose distributions^33,34^ obtained from TPS. Since these previous efforts leveraged heuristic feature engineering, their models are not differentiable and are unable to provide a differentiable map from MLC leaf positions to the probability of PSQA plan failure. This means that their models are not applicable to be directly used in the leaf sequencing algorithms to produce MLC positions that are likely to pass PSQA.

In this study, we develop a tabular transformer neural network model FT-Transformer^35^ based directly on MLC leaf positions to predict volumetric arc therapy (VMAT) PSQA failure. Using 968 patient plans previously treated with 2–4 VMAT arcs, we trained a regression model to predict the ArcCheck-based PSQA gamma pass rates. We evaluated our model in both the regression context and additionally in the classification context of predicting the pass or fail of PSQA by directly computing receiver operating characteristic (ROC) area under the curve (AUC) on the regression predictions.

We compared the performance of our model with the results from two leading gradient boosted decision tree models in their CatBoost and XGBoost implementations^36,37^ widely used for tabular data as well as to a non-learned complexity metric, mean MLC gap.

Neither FT-Transformer nor CatBoost have been used in the context of PSQA failure prediction. Our proposed approach is distinguished from the previous efforts in that we predict PSQA failure directly from MLC leaf positions and the FT-Transformer model we applied is end-to-end differentiable with no heuristic feature engineering. As the MLC leaf positions are the output of leaf sequencing optimization algorithms, our model could be directly leveraged as a differentiable regularizer to improve the leaf sequencing algorithms to produce deliverable treatment plans (i.e., plans with a lower chance of PSQA failure). This is especially useful for the algorithms that employ gradient-based optimization, some of which are implemented in commercial TPS^4,8^.

## II. Methods

In this section, we describe the pipeline of our study including the description of data collection and processing as well as the models, evaluation metrics and hyperparameter tuning approaches we use. This study was approved by the institutional review board of the University of Washington (STUDY00015736).

### II.A. Data Description

We retrospectively collected DICOM-RT PLAN^38^ files of 968 patients previously treated with 2 – 4 VMAT arcs using Elekta linear accelerators with Agility collimators between January 2019 and August 2021. All plans were designed in Raystation TPS^∗^. PSQA of each plan was done using ArcCHECK^†^ and the gamma analysis of each PSQA used the criteria of 3% dose difference and 3 mm distance-to-agreement (3%/3mm). We excluded stereotactic body radiotherapy (SBRT) patients since our clinic applies different criteria for the gamma analysis with SBRT patients. We constructed a tabular dataset on beam level leveraging the DICOM-RT PLAN^38^ files of the treatment plans to form the samples: for each arc in a treatment plan, we used the leaf and jaw positions of the MLC collimators at each gantry angle.

We aggregated the MLC positions by computing the MLC gap for each leaf-jaw pair at every gantry angle and averaging every 10 neighboring MLC pairs. Additionaly, we averaged the gantry angles over every 8-degree sector. For the labels, we used the ArcCheck-based percentage gamma pass rate of each arc obtained as part of the standard PSQA process in our clinic. To obtain the gamma pass rates, we parsed the ArcCheck-generated PDF reports corresponding to each patient using the `PyPDF2`^‡^ Python package. As the result, we obtained a tabular regression dataset with 360 purely numerical features and 1873 samples.

For our ultimate goal of PSQA failure prediction, we consider the same data in the classification context by thresholding the regression labels and converting them into binary classification labels. We defined the action threshold level in the gamma analysis to be at 95 % as is common in clinical practice^39,40,41^ and obtained binary classification labels (pass or fail) based on this threshold. We reserved 65% of the samples for the training set, 15% for the validation set and 20% for the test set. To pre-process the data, we normalized the features and regression targets by subtracting their mean over the training set and dividing by their standard deviation over the training set.

### II.B. Transformer-based tabular deep learning model

#### Background of machine learning models for tabular data

Gradient boosted decision trees (GBDT)^36,37,42,43^ are the traditionally dominant machine learning approaches for tabular data. These models are commonly used in practice and widely deployed in industry in various domains^44^. Although numerous models have been proposed based on using differentiable ensembles^45,46,47,48,49^, leveraging attention-based transformer neural networks^35,50,51,52,53,54^, as well as other approaches^55,56,57,58,59,60^, recent work on systematic evaluation of deep tabular models^35,44^ shows that there is no universally best model capable of consistently outperforming GBDT. Transformer-based models have been shown to be the strongest competitor of GBDT^35,50,54,61,62^, especially when coupled with a powerful hyperparameter tuning toolkit^35,63^.

#### Tabular transformer model

We employ the recent transformer-based tabular deep learning method FT-Transformer proposed by Gorishniy *et al*.^35^ which has been shown to be the strongest neural network approach in the tabular data domain^35,61^. Additionally, we compare the performance of our model with the gradient boosted decision trees, and we use the popular CatBoost^36^ and XGBoost^37^ packages.

#### Evaluation of model performance

We evaluate the models in the regression context of predicting the gamma pass rates as well as in the classification context of predicting the PSQA plan failures. In the regression context, we use mean absolute error (MAE) and root mean squared error (RMSE) metrics as well as Pearson’s and Spearman’s correlation coefficients between the predictions and the ground truth gamma pass rate values. In the classification context, we use the receiver operating characteristic (ROC) area under the curve (AUC) to evaluate the model performance. We report the beam-level ROC AUC and patient-level ROC AUC. The patient-level predictions and labels are obtained by converting the beam-level predictions and labels such that a plan is labeled as fail if at least one beam in the plan failed QA. In the classification context we also evaluate the performance of a non-learned baseline approach based on the average MLC gap^15^ for comparison.

#### Hyperparameter tuning

We use the Optuna Bayesian optimization toolkit^63^ for hyperparameter tuning. The hyperparameter search spaces for each model are reported in Appendix A. To avoid overfitting, we use early stopping with patience for each model, i.e., we stop training the models if no improvement in the validation score is observed for 30 epochs with FT-Transformer or for 50 boosting rounds with CatBoost and XGBoost.

## III. Results

In this section we present the performance of the FT-Transformer model and compare it to the gradient boosted decision trees as well as to the non-learned mean-MLC-gap complexity metric baseline. We investigate the model performance both on the regression task of predicting the ArcCHECK gamma pass rates and the classification task of predicting the QA failure.

### Regression results

We first present the performance of all models in predicting the gamma pass rates in Table 1. For each model we present four regression performance metrics: mean absolute error (MAE), root mean squared error (RMSE), Pearson’s *r* and Spearman’s *r* correlation coefficients. FT-Transformer offers competitive performance with CatBoost and XGBoost and all models achieve good results, with e.g. MAE of the gamma rate predictions between 1.4% and 1.53%. The MAE, RMSE, Pearson’s *r* and Spearman’s *r* values are consistent and are on the same order with the results of other studies in the literature^21,22,23,28,32^ even though they are not directly comparable given the differences in the experimental setups due to the varying hospital equipment and PSQA processes.

**Table 1:**
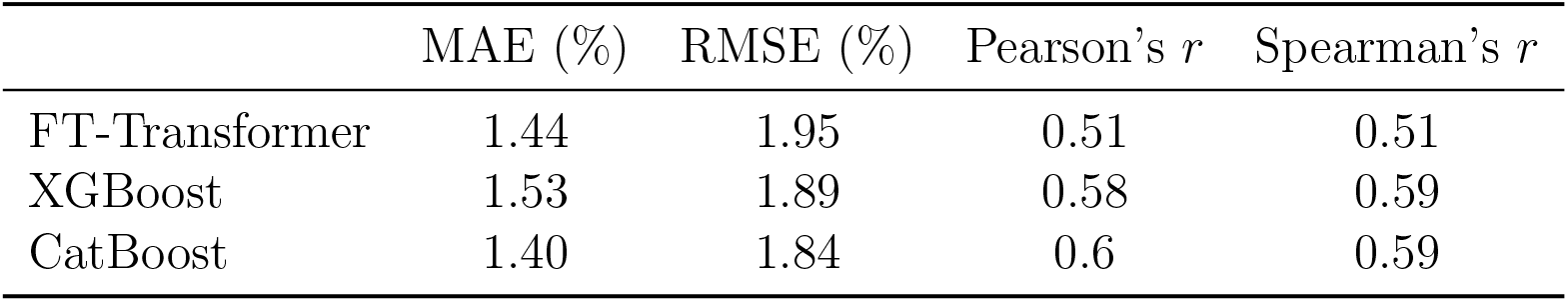
Regression results. Rows correspond to models and columns correspond to regression metrics.

| | MAE (%) | RMSE (%) | Pearson’s $r$ | Spearman’s $r$ |
| --- | --- | --- | --- | --- |
| FT-Transformer | 1.44 | 1.95 | 0.51 | 0.51 |
| XGBoost | 1.53 | 1.89 | 0.58 | 0.59 |
| CatBoost | 1.40 | 1.84 | 0.6 | 0.59 |

### Classification results

The ultimate clinical utility of our models is predicting the PSQA failures to reduce the patient treatment delays and the load on the hospital resources. This practical setup is best emulated by considering our models in the classification context. However, training the models using the regression labels instead of the classification labels directly allows us to leverage more fine-grained target information and avoid the challenges of severe class imbalance in the classification labels. Nonetheless, the predictions of our regression models could be evaluated in the classification context and we present these results in Table 2. We highlight that Table 2 shows two types of ROC AUC metrics: beam-level and patient-level. As mentioned in section II.B., the patient-level predictions are formed from the beam-level predictions by considering a patient plan to be failed if at least one of the beams in the plan is failed.

**Table 2:** Classification results. Rows correspond to models and columns correspond to classification metrics.

|  | Beam-level ROC AUC | Patient-level ROC AUC |
| --- | --- | --- |
| FT-Transformer | 0.82 | 0.85 |
| XGBoost | 0.87 | 0.87 |
| CatBoost | 0.86 | 0.87 |
| Mean MLC Gap Baseline | 0.71 | 0.72 |

As the main takeaways of Table 2, we observe that the patient-level ROC AUC classification performance of FT-Transformer is very close to that of CatBoost and XGBoost and that all of the machine learning approaches significantly outperform the Mean-MLC-Gap baseline.

While ROC AUC summarizes the classification performance for all of the prediction thresholds, a particular threshold has to be selected in practice. To investigate this, we further report the patient-level ROC curves for each of the machine learning models in Figure 1. Since missing a failed plan results in patient rescheduling, it is more costly than sending a successful plan for replanning. Therefore, in our clinical scenario it is beneficial to maximize the true positive rate of PSQA failure identification while keeping the false positive rate at a reasonable value. From the shape of the ROC curves in Figure 1, we observe that FT-Transformer, CatBoost, and XGBoost serve this purpose well and all allow to achieve 80% true positive rate while keeping the false positive rate under 20%.

**Figure 1:**
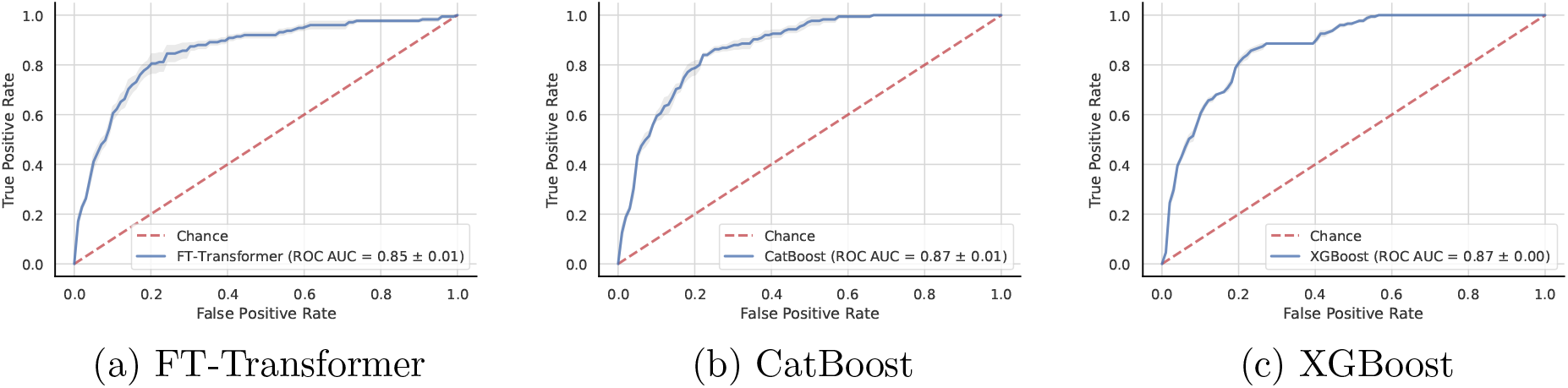
Patient-level ROC curves. (a) FT-Transformer (b) CatBoost (c) XGBoost. The error bars represent the standard error across 5 seeds. The positive label corresponds to plan failure.

## IV. Discussion

We demonstrated that PSQA failure prediction is feasible using just the MLC leaf position data without feature engineering. We evaluated the FT transformer model in both regression and classification contexts and found that it outperforms the non-learned model with a mean MLC gap complexity metric, and performs similarly with the two leading gradient boosted decision tree models, CatBoost and XGBoost. The FT-Transformer neural network model, CatBoost, and XGBoost all provide a substantial improvement over the complexity-metric-based baseline. However, the FT-Transformer model comes with a benefit of being end-to-end differentiable, providing a differentiable map from MLC positions to the probability of PSQA failure. Therefore, this model could be leveraged as a differentiable regularizer that allows gradient-based leaf sequencing optimization algorithms to produce a deliverable treatment plan that is likely to pass PSQA.

It is challenging to directly compare models across different studies due to the lack of existing benchmark datasets and there being numerous combinations of TPS, beam models, linear accelerators, MLC designs, and PSQA procedures, all of which can affect the performance, making apple-to-apple comparison difficult. However, we note that our results are consistent with the performance published in the literature^21,22,23,28,32^. Our models achieve classification performance of 0.85-0.87 ROC AUC and are able to identify 80% of treatment plans that would have failed the PSQA while sending for replanning only up to 20% of successful plans. Using these models in clinical practice can substantially reduce the need for replanning and possibly rescheduling patient due to PSQA failure, which imposes extra workload and stress, and can ultimately compromise patient safety.

Our work was motivated by recognizing the correlation between MLC related complexity metrics and PSQA failures. This leads to the idea of improving leaf sequencing algorithms to produce MLC movements that are more likely to pass PSQA to begin with, which we believe is an improvement from the previous efforts to reduce the frequency of replanning and redoing PSQA by identifying a treatment plan that is likely to fail in the upstream of the workflow, i.e., prior to doing PSQA. We successfully built a model to predict PSQA failure solely based on MLC and jaw positions exploiting recent advances in tabular machine learning models. Incorporating FT-Transformer model in the leaf sequencing algorithms to estimate the potential reduction in the PSQA failure probability of the resulting plans is left for future work.

## V. Conclusion

In this work we applied the leading tabular machine learning approaches to the problem of PSQA failure prediction based solely on MLC leaf positions, and obtained effective models which have both direct clinical practice impact to reduce the PSQA failure as well as potential to improve MLC leaf sequencing algorithms to produce treatment plans that are more likely to pass PSQA.

## Data Availability

The data used in the study is not publicly available

## A Hyperparameter search spaces

### A.1. FT-Transformer

The number of attention heads is always set to 8.

### A.2. Catboost

The hyperparameter search space and distributions are presented in Table 4.

**Table 3:** Optuna hyperparameter search space for FT-Transformer.

| Parameter | Search Space |
| --- | --- |
| Number of layers | UniformInt[1, 4] |
| Feature embedding size | UniformInt[64, 512] |
| Residual dropout | {0, Uniform[0, 0.2]} |
| Attention dropout | Uniform[0, 0.5] |
| FFN dropout | Uniform[0, 0.5] |
| FFN factor | Uniform[2/3, 8/3] |
| Learning rate | LogUniform[$1e-5$, $1e-3$] |
| Weight decay | LogUniform[$1e-6$, $1e-3$] |

**Table 4:** Optuna hyperparameter search space for Catboost.

| Parameter | Search Space |
| --- | --- |
| Max depth | UniformInt[3, 10] |
| Learning rate | LogUniform[$1e-5$, 1] |
| Bagging temperature | Uniform[0, 1] |
| L2 leaf reg | LogUniform[1, 10] |
| Leaf estimation iterations | UniformInt[1, 10] |

### A.3. XGBoost

The hyperparameter search space and distributions are presented in Table 5.

**Table 5:** Optuna hyperparameter search space for XGBoost.

| Parameter | Search Space |
| --- | --- |
| Max depth | UniformInt[3, 10] |
| Min child weight | LogUniform[$1e-8$, $1e5$] |
| Subsample | Uniform[0.5, 1] |
| Learning rate | LogUniform[$1e-5$, 1] |
| Col sample by level | Uniform[0.5, 1] |
| Col sample by tree | Uniform[0.5, 1] |
| Gamma | {0, LogUniform[$1e-8$, $1e2$]} |
| Lambda | {0, LogUniform[$1e-8$, $1e2$]} |
| Alpha | {0, LogUniform[$1e-8$, $1e2$]} |

## Footnotes

∗ RaySearch Laboratories

† Sun Nuclear corporation

‡ https://pypi.org/project/PyPDF2/

